# Awareness and Acceptance of Malaria Vaccines by Caregivers of Under-five Children in Abia State, Nigeria: A Mixed Methods Study

**DOI:** 10.1101/2025.11.17.25340288

**Authors:** Eziyi Iche Kalu, Ugo Uwadiako Enebeli, Benjamin S. Chudi Uzochukwu, Ebelechukwu Lawrence Enebeli, Faith Adamma Kalu, Perfection Chinyere Igwe, Justin Junior Kalu, Beauty Olamma Kalu, Agwu Nkwa Amadi, Yakubu Joel Cherima, Uchenna Stephen Nwokenna, Rejoice Kaka Hassan

## Abstract

**Introduction:** Malaria remains a leading cause of under-five mortality in Nigeria, with Abia State exemplifying hyperendemic transmission. The 2024 introduction of the R21/Matrix-M and RTS,S/AS01 vaccines offers promise, but evidence on caregiver awareness and acceptance in the non-pilot South-East region to inform equitable rollout is scarce.

**Methods:** We conducted a mixed-methods study from June to August 2025 among 618 caregivers of under-fives caregivers engaged with the routine immunization programme in Abia State. Quantitative cross-sectional surveys assessed awareness (knowledge of malaria vaccines) and acceptance (willingness on Likert scales), and inferential analysis was carried out using logistic regression. Qualitative in-depth interviews (n=28) and focus group discussions (n=6, 50 participants) explored perceptions using thematic analysis. Triangulation integrated findings.

**Results:** Awareness was low (38.2%; 95% CI: 34.5-42.0) and highest among urban educated caregivers (52.3%). However, acceptance was high (88.7%), driven by child protection (67.4%) and provider trust (59.8%). Barriers included fears of side effects (51.4%) and misinformation (18.7%). Significantly, education (AOR=3.28) and urban residence (AOR=1.78) predicted awareness, and income (AOR=2.05) and awareness status (AOR=6.92) influenced acceptance. Qualitative themes corroborated quantitative findings: “fragmented information” explained rural gaps, and “maternal instinct” amplified willingness to accept.

**Conclusion:** Based on our findings, caregivers demonstrated strong acceptance of malaria vaccines despite critically low awareness, a disparity fuelled by information gaps and sociodemographic inequities such as low education and rural residence that threaten vaccine rollout and malaria elimination goals. Based on our findings, this pioneering mixed-methods study recommends that specific channels that leverage PHC providers and community leaders for information dissemination should be utilised, given the high levels of trust, to ensure malaria vaccine uptake and accelerate progress in reducing under-five deaths.

**KEY MESSAGES:** *What is already known on this topic:*

- Malaria vaccines (RTS,S/AS01 and R21/Matrix-M) were introduced in Nigeria in late 2024, with prior studies in northern regions reporting low caregiver awareness and high acceptance rates. However, evidence remains scarce in non-pilot South-East states like Abia, where hyperendemic transmission and urban-rural disparities necessitate localised data to guide equitable rollout and integration into routine immunisation.

*What this study adds:*

- This pioneering mixed-methods study in Abia State of Nigeria reveals critically low malaria vaccine awareness (38.2%) among 618 routine immunisation-engaged caregivers, contrasted by robust acceptance (88.7%), with education, urban residence, income, and awareness status as key predictors, corroborated by qualitative themes of “fragmented information” barriers and “maternal instinct” facilitators.
- It provides the first post-approval, pre-expansion insights from Abia State, rejecting the null hypothesis of no sociodemographic associations and highlighting resilience in acceptance despite informational gaps.

*How this study might affect research, practice or policy:*

- Policymakers should integrate these findings into Nigeria’s National Malaria Strategic Plan 2024-2028 with South-East-contextualised, Igbo-language awareness campaigns through church networks and media to bridge the 38.2% awareness gap and align with African Union vaccination targets.
- Programme managers can leverage high provider trust (59.8%) through community health worker-led dialogues and mobile clinics to dispel side-effect myths, targeting low-education rural caregivers and potentially elevating acceptance beyond 88.7% for improved under-five malaria prevention.

**GRAHICAL ABSTRACT:** 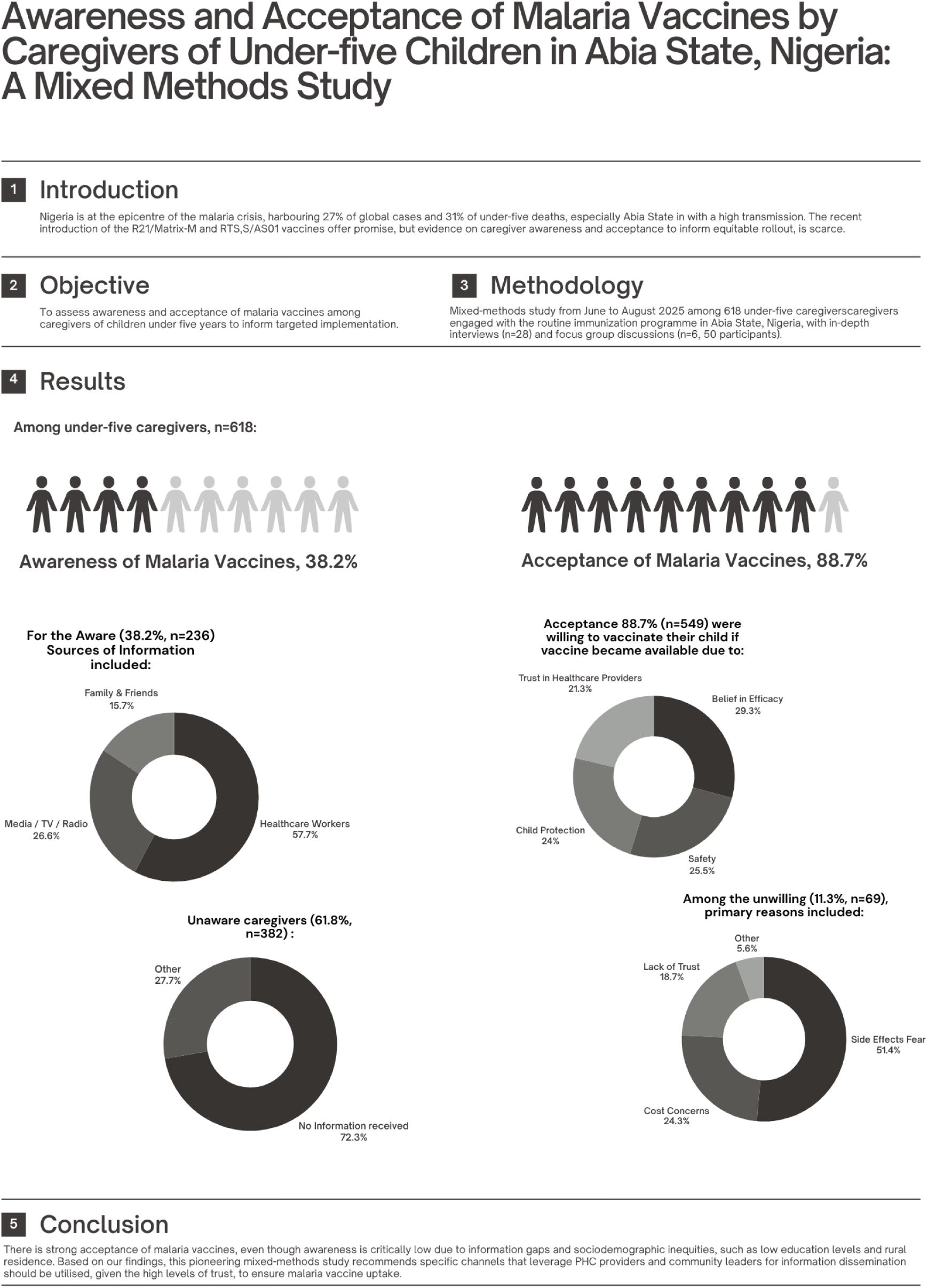

## INTRODUCTION

Malaria remains one of the most devastating infectious diseases globally, with an estimated 263 million cases and 597,000 deaths reported in 2023, representing an 11 million case increase from the previous year.^1^ Sub-Saharan Africa accounts for 94% of cases and 95% of deaths, with children under five years particularly vulnerable.^1^ In Nigeria, the epicentre of the African malaria crisis, the disease constitutes 27% of global cases and 31% of deaths, translating to over 71 million estimated cases annually and an economic toll exceeding $1.1 billion in healthcare costs and productivity losses.^1,2^ Within Nigeria, Abia State in the South-East geopolitical zone exemplifies the region’s disparity, with up to 58.5% malaria prevalence among children under five, driven by ecological factors such as high rainfall, dense vegetation, and urban-rural interfaces that favour *Anopheles* vector proliferation.^3,4^

The advent of malaria vaccines marks a paradigm shift in malaria prevention strategies. The RTS,S/AS01 vaccine, recommended by WHO in 2021 for routine immunisation in moderate-to-high transmission areas, demonstrated a 30% reduction in severe malaria in phase III trials.^5^ Complementing this, the R21/Matrix-M vaccine was prequalified by WHO in 2023 for its high efficacy (in reducing symptomatic malaria cases by 75% during the 12 months following a three-dose series).^6^ Nigeria, aligning with the African Union’s 2023 declaration to vaccinate 18 million children by 2025, approved the R21 vaccine in October 2024 and initiated phased deployment, starting with high-burden states like Bayelsa (south) and Kebbi (north) on December 5, 2024.^7^ The malaria vaccine has been integrated into Nigeria’s routine immunisation schedules (four doses for children aged 5-15 months); however, early studies reveal suboptimal awareness and acceptance, attributed to systemic and behavioural challenges.^8,9^

Awareness and acceptance of malaria vaccines among caregivers in low-resource settings like Nigeria are frequently suboptimal, with logistical hurdles compromising outreach efforts to remote areas.^9^ Hesitancy, fuelled by misinformation, historical distrust in health systems, and misconceptions about vaccine safety, further impedes acceptance. Only 40.3% (northern Nigeria) and 46.1% (Gombe State, Nigeria) of caregivers were aware of the vaccine’s introduction.^8,10^ Socioeconomic factors worsen inequities, particularly among low-income and rural households, whereas facilitators to acceptance include robust community engagement and integration into existing immunisation platforms.^8,9^ Despite these initial insights, evidence from South-East Nigeria, including Abia, where urban commercialisation intersects with rural vulnerabilities, remains scarce, limiting tailored intervention.

This study achieved its objective to address this critical evidence gap as it used a mixed-methods approach to triangulate awareness levels and acceptance perceptions in Abia State. The quantitative component targeted caregivers engaged with the routine immunisation programme at Primary Healthcare Centres (PHCs), a group already interfacing with health services and potentially more receptive to vaccines. While this sampling strategy may introduce selection bias toward higher acceptance rates compared to the broader caregiver population, it enhanced the validity of findings for interpreting uptake potential among this key demographic pivotal to vaccine integration. This study is unique in its timeliness, in its conduct post-malaria vaccine national approval but pre-expansion to South-East states, and offered the first localised, integrated analysis of awareness and acceptance dynamics in a non-pilot, high-burden urban-rural context. This study hypothesised that there was no statistically significant association between sociodemographic factors and malaria vaccine awareness and acceptance in Abia State, Nigeria, and the study’s rejection of this null hypothesis generated actionable, context-specific recommendations to enhance awareness campaigns and acceptance strategies and potentially inform Nigeria’s National Malaria Strategic Plan 2024-2028.

## MATERIALS AND METHODS

### Study Setting

Abia State (coordinates: 4°48’-6°02’N, 7°09’-7°58’E), is located in the South-East geopolitical zone of Nigeria in West Africa and spans an area of approximately 6,320 km² characterised by a tropical rainforest climate with bimodal rainfall patterns that foster perennial mosquito vector breeding sites.^11^ The state is divided into 17 local government areas (LGAs), with a 2025 estimated total population of 4,733,613 persons.^12^

### Study Design

We used a mixed-methods design to allow for triangulation of quantitative data with qualitative depth. The quantitative component was a cross-sectional survey to estimate malaria vaccine awareness levels and associations with sociodemographic factors, while the qualitative component involved in-depth interview (IDIs) and focus group discussions (FGDs) to explore perceptions and contextual nuances. Quantitative and qualitative data were analysed independently before integration, and joint displays integrated the findings.

### Study Participants

The study participants were primary caregivers of children under five years of age in Abia State, Nigeria, attending primary healthcare centres for routine immunisation of their under-five children. These caregivers served as the key respondents in the mixed-methods design, providing insights into awareness and acceptance of the malaria vaccines.

#### Inclusion Criteria

Participants aged ≥18 years or older; Primary caregivers responsible for making healthcare decisions for at least one child under five years; Resident in Abia State for a minimum of six consecutive months prior to data collection; and Willingness to provide informed consent.

#### Exclusion Criteria

Healthcare professionals; Caregivers who were unable to provide informed consent; Caregivers who were too ill to respond; and Caregivers whose children were not under five years of age.

### Sample Size Determination

For the quantitative survey, sample size was calculated using the Fisher’s formula for sample size determination of proportion in health studies:^13^

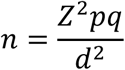

Where: *n* = minimum sample size, Z = 1.96 (95% confidence level), p = 0.403 (malaria vaccine awareness prevalence in northern Nigeria);^10^ q = 1 - p = 0.597; d = 0.05 (desired precision)

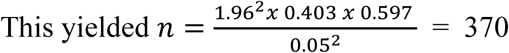

Adjusting for a design effect of 1.5 for multi-stage clustering and a 10% non-response, the minimum sample size was 611 caregivers. We eventually surveyed 618 caregivers.

For the qualitative components, 28 IDIs (14 per urban/rural stratum) and six FGDs (three urban, three rural; eight to nine participants each, n=50 total) were planned, guided by data saturation when no new themes emerge.^14^

### Sampling Technique

A multi-stage sampling technique was used for the quantitative survey. Stage one included the simple random selection of six LGAs (three urban and three rural LGAs) from the 17 LGAs of Abia State. The three urban LGAs were Aba South, Umuahia North, and Osisioma; and the three rural LGAs were Ugwunagbo, Ohafia, and Isiala Ngwa South. Stage two involved simple random selection of two primary healthcare centres (PHCs) per LGA (n=12 PHCs total) using the list of basic healthcare-funded PHCs in Abia State. Stage three entailed systematic random sampling of eligible caregivers attending immunisation clinics based on average daily attendance from clinic registers, for a total of 103 under-five caregivers per PHC.

For qualitative data, purposive sampling targeted information-rich cases: 28 IDI participants were recruited from survey respondents (stratified by low/high awareness scores), ensuring diversity in age, education, and parity. Fifty FGD participants were purposively selected from community leaders’ networks, homogeneous by gender (to encourage openness) and residence.

### Data Collection

Data was collected from June to August 2025, and six trained research assistants (with postgraduate qualifications in public/community health, a minimum of three years of mixed-methods research backgrounds, and fluency in English and Igbo languages) underwent a three-day workshop on protocols, consent administration, and cultural sensitivity. Quantitative surveys were face-to-face, lasted 20-30 minutes, and were conducted in private spaces in the PHCs during routine immunisation sessions with the questionnaire on KoboCollect. Qualitative IDIs (45-60 minutes) and FGDs (60-90 minutes) were held in community halls or homes and audio-recorded with permission using encrypted digital recorders (Olympus WS-882). Field notes captured non-verbal cues. There were daily debriefs, back-end and spot checks to ensure adherence to study protocols.

### Study Instrument

The quantitative instrument was a structured, pre-tested questionnaire adapted from the WHO Vaccine Hesitancy Scale,^15^ and prepared on KoboCollect, comprising four sections: (1) Sociodemographic (included age, education, income, residence, parity; 10 items); (2) Awareness (“informed awareness”) included eight items on ever hearing about malaria vaccine, knowledge of RTS,S/R21 vaccines, vaccine schedule (e.g., four doses, 5-15 months), efficacy, target age group, administration site (intramuscular), and primary information source from which informed awareness was defined as moderate/high score (≥3 correct) to capture meaningful comprehension beyond mere exposure, and unawareness signified a 0-2 score; (3) Acceptance was defined as willing to accept the malaria vaccine for their child when it is available,^10^ and included questions on barriers/facilitators (on 5-point Likert scale; 12 items); and (4) Access perceptions (distance, logistics; five items). The instrument was translated/back-translated into Igbo language and pilot-tested on 65 non-sampled caregivers (Cronbach’s α=0.82) and content validity with five experts. Qualitative guides were informed by the theoretical domains framework,^16^ and were piloted and refined for cultural relevance.

### Data Management

The data on KoboCollect was downloaded as a Microsoft Excel file, and initial cleaning involved range checks and logical consistency (e.g., age ≥18). Files were exported to SPSS for analysis. Qualitative audio files were transcribed verbatim in original languages by certified transcribers, and English translations were quality-checked. Transcripts were uploaded to NVivo 14 for thematic coding, with backups on institutional servers compliant with Nigeria’s Data Protection Act.^17^ Data access was role-based, and audit trails logged all modifications.

### Statistical Analyses

#### Variables

The primary outcomes were malaria vaccine awareness and acceptance among under-five caregivers; predictors were the sociodemographic factors; potential confounders were education, income and residence which were adjusted for; residence and education level were assessed for potential effect modification in the statistical analysis.

#### Analysis

Quantitative analysis used SPSS v29.0. Descriptive statistics summarised awareness (proportions) and acceptance (mean±SD for Likert scores). Bivariate associations utilised chi-square tests/Fisher’s exact for categorical variables and independent t-tests for continuous (p<0.05). Multivariate logistic regression modelled predictors of acceptance (adjusted odds ratios [AORs], 95% CIs), and adjusted for confounders (e.g., education, income).

Subgroup analyses were conducted to explore variations in primary outcomes (awareness and acceptance) across key sociodemographic strata. Interactions were examined through interaction terms in bivariate models, and qualitatively through purposive stratification of IDIs and FGDs to probe contextual nuances. Missing data were addressed using complete case analysis, excluding cases with incomplete responses on primary outcomes. Sensitivity checks confirmed no wide differences in crude awareness (38.2% vs. 38.0%) or acceptance (88.7% vs. 88.5%) when excluding vs. including cases. Qualitative data underwent reflexive thematic analysis in NVivo.^18^ Integration occurred by merging quantitative and qualitative themes, visualised in joint display tables. There were no missing transcripts due to audio-recording protocols and daily debriefs.

### Potential Sources of Bias and Efforts to address them

To address potential selection bias at PHCs, a multi-stage probability sampling was used, while acknowledging potential overestimation of acceptance due to this receptive subgroup. The information bias from self-reporting was mitigated by private interview settings, trained research assistants, and triangulation of quantitative scores with qualitative probes for depth and validation. The exclusion of healthcare professionals minimised response bias from professional knowledge. Confounding was addressed using multivariate logistic regression adjusting for key variables, with bivariate checks for associations.

### Ethical Statement

This study adhered to the principles outlined in the Declaration of Helsinki and was conducted in accordance with ethical guidelines for health research involving human participants. Prior to data collection, ethical approval was obtained from the Ethical and Research Committee of the Abia State Ministry of Health (approval number: AB/MH/PRS/ECS/T.1/1016). And written informed consent was obtained from all participants after a detailed explanation of the study’s purpose, procedures, potential risks (e.g., minimal emotional distress from discussing health concerns), benefits (e.g., contributing to public health policy), and voluntary nature. Participants were informed of their right to withdraw at any stage without repercussion. To ensure confidentiality, all data were anonymised using unique alphanumeric codes and stored on password-protected servers accessible only to the research team.

## RESULTS

A total of 618 caregivers participated in the quantitative survey, achieving a response rate of 98.1% (618/630 approached), with non-response primarily due to time constraints during immunization sessions (n=12 refusals). For the qualitative component, 28 caregivers completed the IDIs, and 50 caregivers participated across six FGDs (8-9 per group). Participant demographics were comparable (e.g., 81.9% female in quantitative vs. 79.0% in qualitative).

### Sociodemographic Characteristics of Participants

The majority of quantitative participants were female (n=506, 81.9%), reflecting primary caregiving roles in the region. Mean age was 32.4±6.2 years (range 18-48), with most (n=381, 61.7%) aged 26-35 years. Over half (n=395, 63.9%) had secondary or higher education, and 52.3% (n=323) resided in urban areas. Household monthly income was low for 44.7% (n=276; <₦30,000 or approximately US$18), and 68.4% (n=423) had one to two under-five children, as in **Table 1**. Qualitative participants mirrored this: 22/28 (78.6%) IDI participants were female, mean age 33.1±5.9 years; FGDs were female-only, with similar education/income distributions.

**Table 1.** Sociodemographic characteristics of quantitative participants (n=618)

| Characteristic | Number, n | Percentage (%) |
| --- | --- | --- |
| **Sex** |  |  |
| Female | 506 | 81.9 |
| Male | 112 | 18.1 |
| **Age group (years)** |  |  |
| 18–25 | 124 | 20.0 |
| 26–35 | 381 | 61.7 |
| ≥36 | 113 | 18.3 |
| **Education level** |  |  |
| No formal/Primary | 223 | 36.0 |
| Secondary | 247 | 40.0 |
| Tertiary or higher | 148 | 24.0 |
| **Residence** |  |  |
| Urban | 323 | 52.3 |
| Rural | 295 | 47.7 |
| **Monthly household income** |  |  |
| <₦30,000 (approximately US\$18) | 276 | 44.7 |
| ₦30,000–60,000 (approximately US\$18–36) | 218 | 35.2 |
| >₦60,000 (approximately US\$36) | 124 | 20.1 |
| **Number of under-five children** |  |  |
| 1–2 | 423 | 68.4 |
| ≥3 | 195 | 31.6 |
| **Employment status** |  |  |
| Employed | 371 | 60.0 |
| Unemployed | 247 | 40.0 |

### Levels of Awareness of the Malaria Vaccine

Overall, 38.2% (n=236/618; 95% CI: 34.5–42.0) of caregivers were aware about the malaria vaccines. Among aware participants, primary sources included healthcare workers (68.4%, n=162), media/TV/radio (31.6%, n=75), and family/friends (18.6%, n=44). Knowledge of vaccine eligibility (children aged 5–15 months in high-transmission areas) was moderate at 54.9% (n=130/236) among aware caregivers, while 41.4% (n=98) correctly identified the four-dose schedule. Unaware caregivers (61.8%, n=382) predominantly cited “no information received” (72.3%, n=276) in open-ended responses.

Figure 1 shows the percentage (95% CI) of awareness of malaria vaccines by educational level (none/primary or secondary+) and residence (rural or urban): Urban Secondary+: 52.3% (45.1-59.5); Urban No/Primary: 28.4% (21.2-35.6); Rural Secondary+: 38.7% (31.4-46.0); Rural No/Primary: 18.9% (13.5-24.3). There was significant urban-education interaction, χ²=42.3, p<0.001).

**Figure 1.**
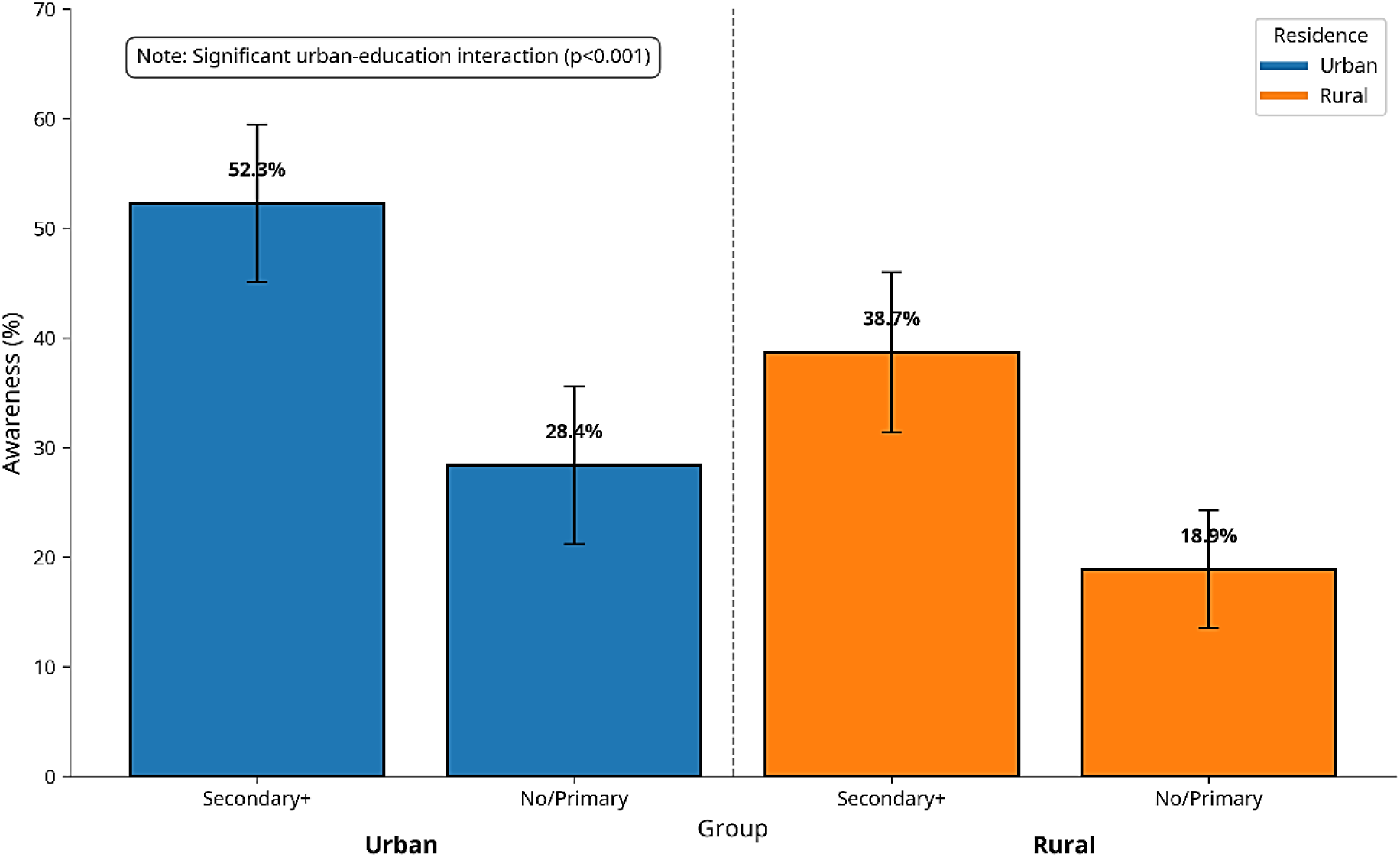
Awareness of malaria vaccine by residence and education level (n=618)

### Qualitative Themes on Awareness of the Malaria Vaccine

The IDI findings from a total of 28 caregivers, and the 50 caregivers across six FGDs aligned with the quantitative data on poor awareness and revealed awareness as a multi-layered construct shaped by incomplete exposure and intrinsic motivations. IDIs and FGDs highlighted sub-themes: “fragmented information” as a barrier to comprehensive understanding, and “maternal instinct” as a facilitator that prompted awareness and information-seeking.

### Sub-theme 1: Fragmented Information

This sub-theme captured the inconsistent, superficial nature of vaccine information reaching caregivers, that failed to translate into actionable knowledge, for example:

“In our village here, nobody is talking about malaria vaccine. I heard it small from one friend of a nurse, but is it for big children or small baby? I don’t know.” (IDI-05, rural female, 26-30 years; reflects 72.3% of unaware caregivers citing “no information received”).

Another rural voice: “I did not hear anything about malaria vaccine. Even when am in the health centre for my baby immunisation, nobody mentions anything.” (FGD-4, rural group, female, 31-35 years; aligns with low rural awareness at 18.9% for low-education subgroups).

Urban FGDs reinforced the theme: “One day they talk about the new vaccine for malaria on the radio, but what is the dose? How many times? We do not hear.” (FGD-3, urban group, female, 31-35 years). These narratives were emergent in 19/28 IDIs and 5/6 FGDs and highlighted how fragmented dissemination perpetuated the 61.8% unawareness rate.

### Sub-theme 2: Maternal Instinct

Contrasting the barriers, maternal instinct (intrinsic motivation) emerged as a driver that compelled caregivers to seek out vaccine information as a foundation for acceptance, despite gaps in information, often framing it as an extension of protective duties.

A semi-urban mother shared: “I first heard about this malaria vaccine from one nurse in a clinic here in Umuahia when I came for immunisation like this. She said its four times from five months and reduce it by 75%. Because malaria is our number one enemy here; any day the vaccine land, I will queue early.” (IDI-09, urban female, 31-35 years; illustrates maternal instinct as a catalyst for awareness by showing how her protective drive against malaria transforms a routine clinic interaction into acceptance of the vaccine).

In FGDs, this instinct amplified collective resolve: “They showed where government was launching it on TV news. One church fellowship sister, her husband is a doctor, he said the two vaccines reduce strong malaria. Well, I don’t know everything, but my child will take it to less malaria suffering when it comes to Abia State.” (FGD-2, urban group, female, 36-40 years; shows how maternal instinct compels her to actively process information from diverse sources and accept the vaccine’s partial efficacy despite incomplete knowledge).

Another IDI captured the emotional urgency: “My sister in Lagos sent me a WhatsApp video about the malaria vaccine, the vaccine will reduce malaria pain, so my child will take it. I plan to ask more about it from health worker another time.” (IDI-23, rural female, 26-30 years; describes an emotional urgency to “reduce malaria pain” in her child which motivates her to seek out further information and awareness).

### Acceptance and Key Perceptions Influencing Acceptance

Malaria vaccine acceptance was high, with 88.7% (n=549/618; 95% CI: 85.9-91.5) of caregivers willing to vaccinate their child when available, including 76.5% (n=473) “very willing” on a 5-point Likert scale (mean=4.2±0.9). Among the unwilling (11.3%, n=69), the primary reasons were side effect fears (51.4%, n=35) and cost concerns (24.3%, n=17). Positive perceptions included belief in efficacy (82.3%, n=509) and safety (71.9%, n=444; mean Likert=3.9±1.0). The facilitators endorsed most were child protection (67.4%, n=417) and trust in healthcare providers (59.8%, n=370).

**Table 2** shows a breakdown of acceptance drivers and barriers, responses to the 12 Likert-scale items (1=Strongly Disagree to 5=Strongly Agree), with means±SD and percentage agreement (% Agree/Strongly Agree). Items were grouped into facilitators (n=6) and barriers (n=6) for clarity. Overall, facilitator items showed higher endorsement (mean=3.8±0.8) compared to barriers (mean=2.4±1.1; p<0.001), supporting the high acceptance rate.

**Table 2.** Detailed Likert-Scale Responses on Drivers and Barriers to Malaria Vaccine Acceptance (n=618)

| Item No. | Item Description | Mean ± SD | Agree /Strongly Agree, n (%) | Category |
| --- | --- | --- | --- | --- |
| **Facilitators** |  |  |  |  |
| 1 | The malaria vaccine will protect my child from severe malaria episodes. | 4.3 ± 0.7 | 551 (89.2%) | Efficacy |
| 2 | I trust healthcare providers who recommend the malaria vaccine. | 4.1 ± 0.8 | 509 (82.4%) | Trust |
| 3 | If the vaccine is free and part of routine immunization, it will make it accessible. | 4.0 ± 0.9 | 486 (78.6%) | Accessibility |
| 4 | If community members vaccinate their children, I am more likely to do so. | 3.9 ± 1.0 | 447 (72.3%) | Social Influence |
| 5 | The vaccine will reduce my family’s healthcare costs and hospital visits. | 3.8 ± 0.9 | 427 (69.1%) | Economic Benefit |
| 6 | WHO approval reassures me of the vaccine’s safety and effectiveness. | 3.7 ± 1.0 | 404 (65.4%) | Credibility |
| **Overall Facilitator Mean:** |  | **3.8 ± 0.8** | **470 (76.2%)** |  |
| **Barriers** |  |  |  |  |
| 7 | I worry about side effects like fever or swelling after vaccination. | 3.2 ± 1.2 | 318 (51.4%) | Safety Concerns |
| 8 | Long-term effects (e.g., infertility) make me hesitant about the vaccine. | 2.8 ± 1.3 | 239 (38.7%) | Misinformation |
| 9 | The distance to the health facility is a major barrier to getting the vaccine. | 2.6 ± 1.1 | 200 (32.4%) | Logistics |
| 10 | Even if free, transport costs to the clinic are too high for my family. | 2.5 ± 1.2 | 180 (29.1%) | Economic Barrier |
| 11 | Misinformation from social media or rumours discourages vaccine uptake. | 2.4 ± 1.0 | 152 (24.6%) | Distrust |
| 12 | Past experiences with government vaccine programs make me sceptical. | 2.3 ± 1.1 | 135 (21.8%) | Historical Distrust |
| **Overall Barrier Mean** |  | **2.6 ± 1.1** | **204 (33.0%)** |  |

Likert scale: 1=Strongly Disagree, 4=Agree, and 5=Strongly Agree

**Table 3.**
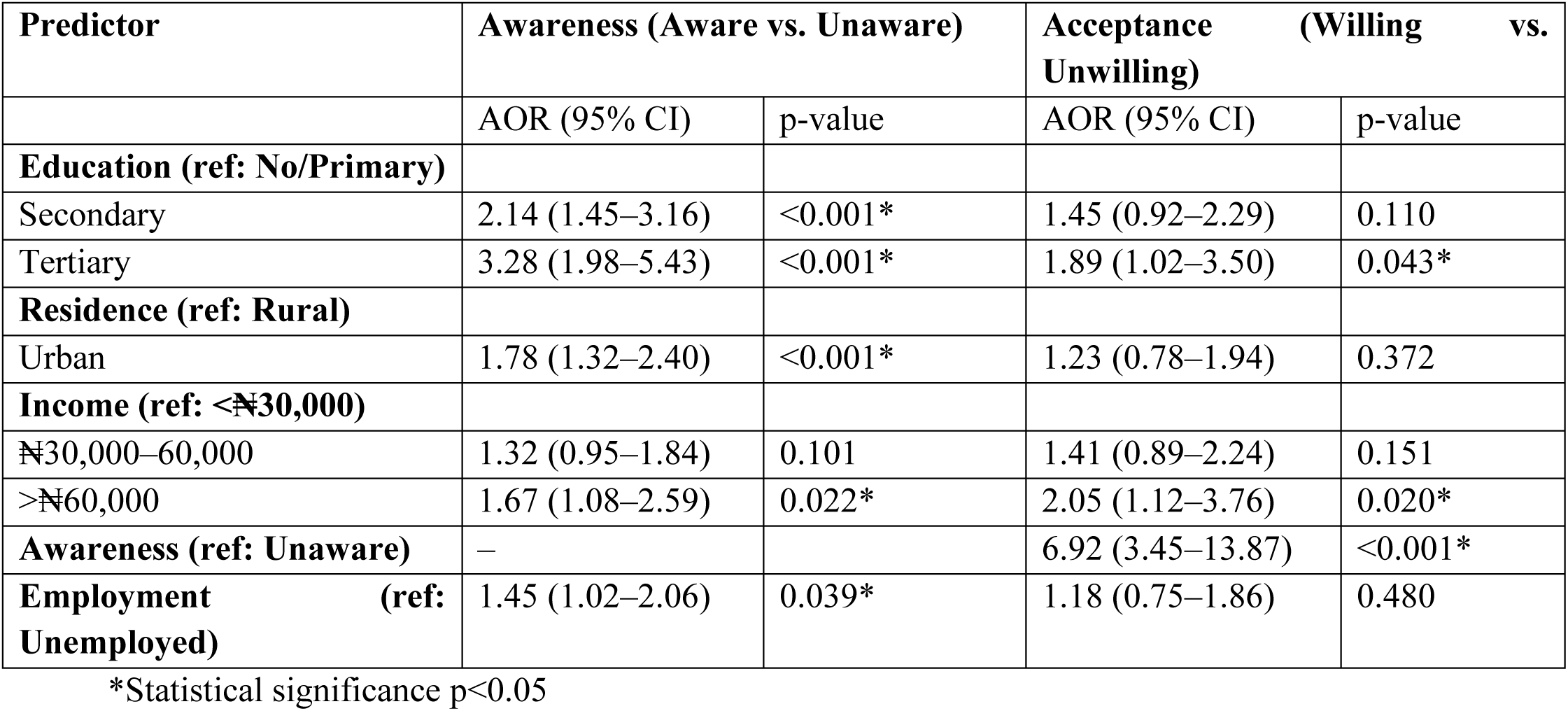
Predictors of Malaria Vaccine Awareness and Acceptance (n=618)

| Predictor | Awareness (Aware vs. Unaware) |  | Acceptance (Willing vs. Unwilling) |  |
| --- | --- | --- | --- | --- |
|  | AOR (95% CI) | p-value | AOR (95% CI) | p-value |
| **Education (ref: No/Primary)** |  |  |  |  |
| Secondary | 2.14 (1.45–3.16) | <0.001* | 1.45 (0.92–2.29) | 0.110 |
| Tertiary | 3.28 (1.98–5.43) | <0.001* | 1.89 (1.02–3.50) | 0.043* |
| **Residence (ref: Rural)** |  |  |  |  |
| Urban | 1.78 (1.32–2.40) | <0.001* | 1.23 (0.78–1.94) | 0.372 |
| **Income (ref: <N30,000)** |  |  |  |  |
| N30,000–60,000 | 1.32 (0.95–1.84) | 0.101 | 1.41 (0.89–2.24) | 0.151 |
| >N60,000 | 1.67 (1.08–2.59) | 0.022* | 2.05 (1.12–3.76) | 0.020* |
| **Awareness (ref: Unaware)** | – |  | 6.92 (3.45–13.87) | <0.001* |
| **Employment (ref: Unemployed)** | 1.45 (1.02–2.06) | 0.039* | 1.18 (0.75–1.86) | 0.480 |

\*Statistical significance p<0.05

Qualitative triangulation highlighted convergence/divergence: The quantitative fears of side effects (51.4%) aligned with IDI themes of “injection risks”, and FGDs added sociocultural layers like spousal influence (absent in quantitative findings, e.g., “Husband must agree, he fears new things” - FGD-5, rural). Facilitators like trust (59.8%) were amplified in qualitative findings as “health worker endorsement” (e.g., “If nurse says safe, I believe” - FGD-2, urban).

### Qualitative Themes on Acceptance of the Malaria Vaccine

Acceptance was predominantly positive (88.7% willingness), framed by “child protection priority” and trust, but tempered by hesitancy from “injection risks” and rumours. Qualitative insights revealed emotional layers where maternal instincts drove uptake, and distrust (e.g., from past vaccine rumours) fuelled reluctance. The following statements exemplify pro-acceptance views (e.g., efficacy, equity) and anti-acceptance concerns (e.g., safety fears).

### Sub-theme: Positive Perceptions and Willingness (Facilitators)

“I will accept this vaccine 100%. My last born suffered serious malaria three times last year, plus the hospital bill chop my small teacher salary. If this malaria vaccine will give my son some protection like they said, and it’s free oh, why not?” (IDI-12, urban female, 31-35 years; echoes 67.4% endorsing child protection and 59.8% trust in providers).

“Acceptance? Yes o. Malaria is like a thief that is stealing our children’s joy, shorting their blood, making them lose weight, and miss school. In my group, we all agree: if health worker agrees with it, it works for us. Because it will even help us to save money for food for family. Also, no cultural things hold me back.” (FGD-3, rural group, female, 36-40 years; aligning with 82.3% belief in efficacy and economic benefits reported in similar African contexts).

“I accept the vaccine because it does not have serious severe side effect like some rumours talk. My friend baby took it in another state, nothing do am.” (IDI-07, rural female, 26-30 years; reflects 71.9% safety perceptions and peer influence in 32.6%).

### Sub-theme: Negative Perceptions and Hesitancy (Barriers)

“Against? Yes, me I no sure. Last time with Penta vaccine, my son leg swell big-big. Now this malaria one, they even say four-four dose, what if it cause infertility or weak blood? They even say it does not work 100%, so like us with road bad, clinic far; if side effect come, who will rush the child back to the health centre? Well, me I will wait for my neighbour to try first before I consider.” (IDI-16, rural female, 31-35 years, primary education; captures 51.4% fear of side effects and 24.3% cost/access concerns).

“I will not accept easy-easy because government is no trustworthy again. Remember that rumour about vaccine that will make woman no born? My husband even hear from his brother that malaria vaccine is experiment on our children.” (FGD-6, rural group, female, 41-45 years; illustrates 18.7% misinformation/distrust, amplified by spousal influence in 26.8% unwilling cases).

“Hmm, cost and distance are a problem. Because, even if they say it is free, transport to health centre is ₦2,000; and if queue is too long or vaccine finish, it becomes a total waste. In my culture too, all these foreign medicines are becoming too much. In short, I will accept only if I see many other children take it first, not my daughter first.” (IDI-25, urban low-income female, 26-30 years; ties to 24.3% affordability barriers and cultural hesitancy in qualitative insights).

### Associations Between Sociodemographic Factors and Awareness/Acceptance

Bivariate analyses showed significant associations between awareness and education (χ²=28.4, p<0.001), residence (χ²=15.7, p<0.001), income (χ²=12.3, p=0.002), and employment (χ²=9.8, p=0.002); no associations with sex, age, or parity (p>0.05). For acceptance, associations were with education (χ²=22.1, p<0.001), income (χ²=18.6, p<0.001), and awareness status (χ²=45.2, p<0.001).

Multivariate logistic regression showed that higher education (secondary: AOR=2.14, 95% CI: 1.45-3.16; tertiary: AOR=3.28, 95% CI: 1.98–5.43) and urban residence (AOR=1.78, 95% CI: 1.32-2.40) predicted awareness. For acceptance, awareness (AOR=6.92, 95% CI: 3.45-13.87), higher income (>₦60,000: AOR=2.05, 95% CI: 1.12-3.76), and education (tertiary: AOR=1.89, 95% CI: 1.02-3.50) were key predictors.

Qualitative data supported higher educated urban caregivers describing “access to information on phones” (e.g., IDI-15), while low-income rural voices highlighted barriers (FGD-6).

These findings thus provide evidence to reject the null hypothesis Ho, that there were no statistically significant associations between sociodemographic factors and malaria vaccine awareness and acceptance.

## DISCUSSION

This mixed-methods study provided the first localised evidence on awareness and acceptance of malaria vaccines (RTS,S/AS01 and R21/Matrix-M) among caregivers of under-five children in Abia State, South-East Nigeria, a hyperendemic region that is not part of the initial national rollout states. Our findings revealed suboptimal awareness (38.2%) but with high acceptance (88.7%), influenced by sociodemographic factors such as education, residence, and income. Qualitative insights highlighted fragmented information channels as a primary barrier to awareness, while child protection desires and trust in healthcare providers drove acceptance, tempered by fears of side effects and misinformation. Our results aligned with other settings in some areas, but are divergent in other areas, highlighting the need for context-specific interventions to bridge the awareness-acceptance gap and accelerate equitable vaccine integration into routine immunisation.

### Awareness of Malaria Vaccines

The low awareness level (38.2%; 95% CI: 34.5-42.0) in our study in Abia State reflected systemic dissemination challenges, but a single-centre study in Northcentral Nigeria had a lower (23.7%) acceptance.^19^ Our finding however, approximated the 40.6% awareness in Northern Nigeria;^10^ but was lower than the 46.1% awareness among caregivers in Gombe State,^8^ which may be attributed in part to proximity to rollout sites and targeted radio campaigns. Our findings on level of awareness were also lower than among women in Ghana’s 2022 malaria indicator survey with awareness from 46.5% to 61.5%.^20^ Qualitative themes of “limited exposure” (e.g., “information did not reach the village”) triangulate with these disparities, echoing the northern Nigeria’s findings of 72.3% unaware caregivers citing “no information,” ^8^ and emphasising the need for South-East-specific channels like Igbo-language radio and church networks.

### Acceptance of Malaria Vaccines

Despite the low malaria vaccine awareness in this study, the acceptance was robust at 88.7% (95% CI: 85.9-91.5), aligning with the global norm that acceptance often outpaces knowledge in novel vaccines.^21^ This enthusiasm was explained as due to “child protection priority” among caregivers in our study. The 88.7% malaria vaccine acceptance in the index study mirrored the 91.9% acceptance in Northern Nigeria,^10^ and the 90.6% acceptance in Burundi;^22^ but was lower than the 97.3% acceptance in North-Central Nigeria;^19^ and higher than the reported acceptance rate of 78.4% for Nigeria, a disparity possible from the minimal national sample size of 347 by Emmanuel et al.,^23^ compared to the robust sub-national sample sizes of 618 in this study and 504 in Ajayi et al.^10^’s study, and may have affected Emmanuel et al.^23^’s study’s generalisability.

The drivers of hesitancy in this study included side effects (51.4%), cost (24.3%), and distrust (18.7%) which were less pronounced in Abia than in Gombe State, Nigeria, with 68% having fears of side-effects in Gombe State.^8^ Narratives like “wait and see the neighbour try first”, as in this study, echo global vaccine hesitancy scales,^24^ Yet Abia’s high baseline acceptance suggested that targeted debunking could yield higher acceptance rates.

Our study reported a striking disparity between low awareness (38.2%) and high acceptance (88.7%) in Abia State, with an underlying resilience in caregiver decision-making that superseded informational deficits, a phenomenon observed in other low-resource settings where behavioural dynamics outweighed comprehensive knowledge.^25–28^ We hypothesize that provider trust (59.8% endorsement) acts as a potent cognitive shortcut, enabling caregivers to extrapolate partial or hearsay information and interpersonal endorsement from familiar health workers into confident action, as evidenced in qualitative narratives like “If nurse says safe, I believe” (FGD-2), which bypassed the need for detailed self-education by leveraging established relational bonds forged through routine immunization interactions.^29^ Similarly, maternal instinct, which manifested as an innate protective imperative, amplified receptivity to even fragmented vaccine information, prioritizing child welfare over evidentiary gaps, as quotes like “As mama, I no fit watch my baby suffer serious fever again” (IDI-09) illustrate how evolutionary drives to safeguard offspring convert vague awareness into urgent intent, aligning with psychological models positing that parental motivation buffers against knowledge barriers in paediatric health behaviours.^30^ These mechanisms imply transformative potential for health communication strategies: rather than exhaustive didactic campaigns, interventions should harness trust through community health worker-facilitated dialogues and instinct through narrative storytelling (e.g., peer testimonials emphasising protection), potentially elevating informed awareness while sustaining acceptance rates.

### Associations Between Sociodemographic Factors and Awareness/Acceptance

In this study, multivariate analyses associated education (AOR=3.28 for tertiary, 95% CI: 1.98-5.43) and urban residence (AOR=1.78, 95% CI: 1.32-2.40) as key predictors of awareness and aligned with Northern Nigeria’s findings, where the level of education (AOR; 0.42; CI 0.23–0.78) predicted acceptance.^10^ Nationally, Isah et al.^9^ found that urban residence was associated with awareness across Nigeria and attributed the rural deficits in awareness to infrastructural barriers, such as limited media penetration.

For acceptance, the awareness status (AOR=6.92, 95% CI: 3.45-13.87), higher income (AOR=2.05, 95% CI: 1.12-3.76), and tertiary education (AOR=1.89, 95% CI: 1.02-3.50) emerged as predictors, consistent with Ajayi et al.^10^’s report that awareness status (AOR = 6.88; 95% CI 1.53–30.99) significantly predicted vaccine acceptability. Similarly, the systematic review of 18 studies reported that lower socioeconomic status was significantly associated with decreased acceptance (OR=0.18, 95% CI, 0.02-0.38).^31^ Globally, MacDonald et al.^21^ highlighted awareness as a universal moderator (pooled OR=5.2, 95% CI: 4.1-6.6), with socioeconomic gradients persisting post-education in RTS,S trials.^32^

Qualitative triangulation reinforced these findings, as educated urban caregivers described “access to information on phones” (IDI-15), and low-income rural barriers (as in FGD-6), approximated Bushi et al.^31^’s observed transport inequities.

### Implications for Policy and Practice

This study’s findings advocate for tailored strategies for malaria vaccines including (a) Awareness amplification through community health worker-led Igbo/Pidgin multimedia (e.g., radio jingles, WhatsApp videos), targeting rural low-education gaps, and integrating with existing structures where feasible; ^33,34^ (b) Hesitancy mitigation through community dialogues addressing side-effect myths, leveraging the provider trust to improve acceptance; and (c) Equity measures like mobile clinics for low-income/rural access, informed by geospatial mapping.

### Limitations and Future Directions

The study’s self-reported data may inflate malaria vaccine acceptance due to social desirability, although triangulation with qualitative probes mitigated this effect. The cross-sectional design cannot establish causality; therefore, longitudinal tracking of caregivers and their under-fives after rollout is recommended. Future research should utilise community-based surveys, evaluate the impacts of interventions, include male caregivers for gender dynamics, and extend the study to other states.

## CONCLUSION

In Abia State, Nigeria, a sentinel of Africa’s malaria hyperendemicity, the routine immunisation-engaged caregivers exhibited a contrast of profound acceptance of malaria vaccines amid alarmingly low awareness, which is generalisable to similar hyperendemic contexts in Nigeria and other sub-Saharan regions. This result is limited by a potential selection bias toward a receptive subgroup; however, it highlights a vaccine readiness eclipsed by informational gaps that risk derailing the nation’s ambitious rollout of RTS,S/AS01 and R21/Matrix-M. Sociodemographic inequities, particularly low/no education and rural residence, exacerbate these disparities, yet qualitative narratives of maternal imperatives and provider trust present green areas for rapid amplification. This mixed-methods evidence is one of the first from the South-East of Nigeria, and signals that bridging the awareness gap through targeted dissemination, leveraging trusted PHC providers for clinic-based education and community leaders for village dialogues, could propel uptake beyond continental benchmarks, avert under-five deaths annually, and fortify Nigeria’s trajectory towards the African Union’s 2025 vaccination target.

## DECLARATIONS

### Author contributions

All authors have contributed to the study. EIK: conceptualisation, data collection, data analysis, review, writing original draft, and funding. UUE: conceptualisation, methodology, analysis, review, editing, supervision, writing final draft. BSCU: supervision, methodology, project administration, editing, review. ELE: software, visualisation, data analysis, editing. FAK: formal analysis, validation, review, editing. PCI: investigation, resources, data curation, review. JJK: methodology, review, editing, funding. BOK: formal analysis, validation, review, editing. ANA: investigation, editing, supervision, review, writing final draft. YJC: formal analysis, validation, review, editing, project administration. USN: methodology, editing, review, project administration, formal analysis. RKH: editing, review, validation, writing final copy. All authors approved the final version of the manuscript, and EIK is the guarantor. The corresponding author attests that all listed authors meet authorship criteria and that no others meeting the criteria have been omitted.

### Funding Statement

This research received no specific grant from any funding agency in the public, commercial or not-for-profit sectors.

### Competing interests

None declared.

### Patient And Public Involvement

Patients and/or the public were involved in the design, or conduct, or reporting, or dissemination plans of this research. Refer to the Methods section for further details.

### Data Availability Statement

The dataset is available from the corresponding author on reasonable request.

### Ethics Statement

Ethical approval was obtained from the Ethical and Research Committee of the Abia State Ministry of Health (approval number: AB/MH/PRS/ECS/T.1/1016). Participants gave informed consent to participate in the study before taking part.

### Consent for Publication

Not applicable. This manuscript does not contain any individual person’s data in any form (including individual details, images, or videos). All data are presented in aggregate form only.

## Acknowledgments

We thank the under-five caregivers for their participation in this study, and the six research assistants on the study for their assistance with data collection.

## Notes

### Competing Interest Statement

The authors have declared no competing interest.

